# Body Composition Changes with Semaglutide: A Systematic Review and Meta-Analysis

**DOI:** 10.1101/2025.09.29.25336760

**Authors:** Guilherme Giorelli, Milton Mizumoto, Silvia Sartoretto, Socorro Giorelli, Paulo Giorelli, Alessandra Bedin-Pochini, Diogo Toledo, Gabriel Barreto, Bryan Saunders

**Affiliations:** Hospital Israelita Albert Einstein, São Paulo, Brasil; Nutrology Academy, Rio de Janeiro, Brasil; Population Health Sciences, Bristol Medical School, University of Bristol, Bristol, UK; Applied Physiology and Nutrition Research Group – School of Physical Education and Sport and Faculdade de Medicina FMUSP, Universidade de São Paulo, São Paulo, Brazil; Center of Lifestyle Medicine, Faculdade de Medicina FMUSP, Universidade de São Paulo, São Paulo, Brazil

## Abstract

**Introduction:** Semaglutide is an effective intervention for weight loss, but the relative contributions of fat and fat-free mass (FFM) to total weight loss remain unclear.

**Methods:** This was a systematic review and meta-analysis of randomized controlled trials (RCTs) evaluating semaglutide use in adults living with overweight or obesity. Searches were performed in PubMed, Web of Science, Embase, and Scopus through April 2025. Eligible trials reported baseline and post-treatment measures of body mass, fat mass, and FFM. Data extraction followed PRISMA guidelines, and risk of bias and certainty of evidence were assessed. Random-effects models were applied, with linear or quadratic functions fitted as appropriate.

**Results:** Seven RCTs involving 452 participants (259 semaglutide, 193 placebo) met the inclusion criteria. Semaglutide produced significant non-linear weight loss (0.04 kg/day; 95% CI 0.03–0.05; p<0.0001), primarily due to fat mass reduction (0.03 kg/day; 95% CI 0.01–0.05; p=0.007). FFM declined linearly at a smaller rate (0.007 kg/day; 95% CI 0.003–0.010; p=0.0001). Certainty of evidence was moderate.

**Conclusion:** Semaglutide induces substantial reductions in body and fat mass, with modest FFM loss. Clinical strategies combining semaglutide with resistance exercise and nutritional support may help preserve lean mass and optimize long-term outcomes.

## Introduction

Obesity is a growing global health crisis^1^ and a major risk factor for chronic diseases such as type 2 diabetes, cardiovascular disease, and certain cancers^2,3^. Traditional weight management strategies, such as dietary modifications, increased physical activity, and behavioural interventions, often yield limited long-term success. Consequently, there has been a growing focus on pharmacological interventions to aid in weight loss^4^, leading to the development of novel medications that target key physiological mechanisms regulating food intake, metabolism, and energy balance.

Semaglutide, a glucagon-like peptide-1 (GLP-1) receptor agonist, has been shown to be an effective strategy to reduce body weight in individuals living with obesity^5^. Several meta-analyses have shown long-term decreases in weight loss of approximately 12%^6-8^. Recent redefinitions of obesity have emphasised that the condition is not solely excess body weight or BMI, but rather the presence, distribution, and function of adipose tissue and its impact on health^9^. In this context, although semaglutide has been shown to be an effective drug intervention for weight loss, the quality of weight loss is important, as preserving muscle mass is critical for metabolic health and long-term outcomes^10^. Evidence regarding the effect of semaglutide on body composition appear mixed, with some studies reporting minimal loss of lean body mass and others suggesting more substantial reductions^11,12^. This highlights the importance of evaluating changes in body fat and fat-free mass, rather than body weight alone, when assessing the therapeutic benefits of semaglutide.

The aim of this study was to conduct a systematic review and meta-analysis of randomized controlled trials to quantify the effects of semaglutide treatment on total body mass, fat mass, and fat-free mass. By distinguishing between fat and fat-free mass loss, this analysis aimed to clarify the composition of body mass loss induced by semaglutide and inform clinical practice to optimize body composition and preserve metabolic health.

## Methods

This study was pre-registered in PROSPERO in July of 2024 (CRD42024564458). The study protocol was designed in accordance with PRISMA guidelines ^13^ and inclusion criteria defined according to PICOS criteria (Population, Intervention, Comparator, Outcomes and Study design). The ***Population*** stipulated that participants must be humans living with overweight (BMI >25 kg/m2) or obesity (BMI >30 kg/m2) with or without any comorbidities. The ***Intervention*** required any form of Semaglutide application at any dose for any duration, with a placebo or non-placebo control (*i*.*e*., no semaglutide received) as a ***Comparator***. Obligatory ***Outcomes*** included body mass, fat mass and lean, fat-free or muscle mass as measured by any means (*e*.*g*., bioimpedance, dual-energy X-ray absorptiometry or other) at pre-application (Baseline) and any moment post-application (of both semaglutide and placebo/control). Included ***Study designs*** were RCTs with a parallel group or crossover design.

### Search strategy

Only English-language peer-reviewed, original human studies were included within this review. An electronic search of the literature was undertaken using PubMed, Web of Science, Embase and Scopus to identify relevant articles using the following search terms: (Ozempic OR Semaglutide) AND (Weight OR Fat OR Muscle OR Lean mass). No date limit was set, and the initial search occurred in July of 2024. An example search strategy is included as a Supporting information (Figure S1 in Supporting information). An updated search was performed in April of 2025.

### Screening

All duplicates were removed before a two-phase search strategy was performed independently by two researchers (GB and BS). At phase one, the researchers were assisted by a large language model (LLM – OpenAI API in RStudio; gpt-4-turbo model; *create_chat_completion* function with temperature 0.1), which classified each abstract according to the previously described PICOS criteria. All abstracts were independently screened by both researchers in a blinded manner. Final decisions were made jointly, following a predefined order of relevance from those meeting all five criteria to those meeting none. Studies with uncertain suitability, including those without specific information as to whether body composition measurements were taken, were maintained at this stage of the search and a final decision was reached at the next phase to ensure that all available data was obtained. However, post-hoc analysis studies were removed in phase one. In phase two, full articles were retrieved and assessed against the eligibility criteria.

### Data extraction

Data extraction was conducted by BS using a standardised extraction spreadsheet. The following information was extracted from each article: Authors & year of publication; Participants (number, age, sex, comorbidities); Semaglutide protocol (method of administration, dose, duration); Placebo information (compound; method of administration, dose, duration); Measurements (body mass, fat mass, muscle mass or lean mass or fat free mass [At multiple time points]; analysis method); information on concurrent interventions (diet [Y/N]; exercise [Y/N]; specifics). Where possible, individual participant data was extracted. All extracted data are available in Table S2 in Supporting information.

An email request to provide data was sent to the corresponding authors via email when data were not available either in text or in figures. Since individual data was not reported in any study, an email request to provide data was sent to all corresponding authors. If no immediate response was received, up to two follow-up emails were sent interspaced by at least two weeks. One study provided anonymised individual participant data directly upon contact ^14^, while individual participant data from Heise et al. ^15^ was made available through the Vivli platform (https://vivli.org/) ^16^. Hansen et al. ^17^ provided means and standard deviations of requested data for all measured time points. Individual data for three studies ^5,11,12^ was not shared with the cited reason being due to ongoing internal assessment for a data sharing platform, but means and standard deviations were made available through redacted clinical study reports. Other individual data requests received non-responses upon multiple contacts ^18^, but median changes and interquartile ranges were available in appendices and subsequently converted for analyses (see statistics section).

### Risk of bias (ROB2) and Grading of Recommendations, Assessment, Development and Evaluations (GRADE)

The three main outcomes (body mass; fat mass; fat-free mass) were graded according to the Grading of Recommendations, Assessment, Development and Evaluations (GRADE) framework ^19^. According to the framework, the certainty of evidence could be considered “high”, “moderate”, “low”, or “very low” depending on the number of downgrades that were attributed to each of the following five topics: 1) Risk of bias, 2) Imprecision, 3) Inconsistency, 4) Indirectness, and 5) Publication bias. <u>Risk of bias</u> was assessed by one author (BS) using the revised tool for assessing risk of bias in randomized parallel-group and crossover trials ^20^. Risk of bias was judged to be ‘Low’ if all domains were considered low risk; ‘Some concerns’ if at least one domain had ‘some concerns’ and ‘High risk of bias’ if at least one domain was at high risk or >3 domains had ‘some concerns’ ^20^. <u>Imprecision</u> was deemed to be present if decision making would be different when the lower and upper confidence limits (95% CIs) were considered as the real effect or if outcomes were calculated from only a few studies with small sample sizes. <u>Inconsistency</u> was determined according to heterogeneity measures (I^2^ and tau^2^). If the study population from the included studies differed from the target population (individuals with obesity), then certainty was downgraded due to <u>Indirectness</u>. Funnel plots were visually inspected to assess for <u>Publication bias</u>. Outcomes were upgraded in the presence of either: 1) a large magnitude of effect, 2) a dose-response gradient or 3) an indication that confounding factors would likely reduce rather than increase magnitude of the effect.

### Statistical Analyses

All analyses were performed with R (version 4.1.1) using the *metafor* package. Data was obtained from studies as change scores and standard deviations (SDs). When individual data was available ^14,15^, these were calculated by averaging individual patient delta and obtaining its SDs. Mean differences (MDs) and standard errors (SEs) were calculated using these change scores and SDs based on the formula by Borenstein ^21^ assuming unequal variances (heteroscedasticity) between groups. When only median differences and interquartile range was available ^18^, they were converted into mean and SD with the formula suggested in Wan, Wang, Liu and Tong ^22^. For the analysis of body mass changes, a mixed-effects (time as fixed, and time nested within study ID as random effect) meta-analysis with meta-regression was performed with treatment time as a quadratic polynomial model (time + time^2^) due to apparent non-linear effects. A similar quadratic model was done for fat mass, though no random effects were included (single time points). An attempt to fit a quadratic model for fat-free mass was also done, and a simple linear fixed-effects had a better fit. For all models, the intercept was set to zero due to the expected zero response at 0 treatment days. The I^2^ was calculated as a measure of heterogeneity between studies for the fixed-effects model. Sigma^2^ and tau^2^ values are also reported in this case. Funnel plots were created with the ‘funnel’ function for visual evidence of asymmetry and potential publication bias.

## Results

### Search results

The initial search retrieved 6528 results across all databases (Figure 1). Duplicates were removed using the Rayyan automation tool which detected 4853 duplicates that were excluded, leaving 3489 potential studies remaining. Following title and abstract evaluation, 109 studies remained for full analysis, of which six attained the inclusion requirements. The updated search retrieved 3547 results across all databases. A total of 2132 duplicates were removed, leaving 1525 potential studies remaining. Following title and abstract evaluation, 19 studies remained for full analysis, of which one achieved the inclusion requirements. The flowchart for screening of both searches is reported in Figure 1. Seven studies totalling data from 452 participants (259 receiving semaglutide, 193 placebo) were included in the analyses (Table S3).

**Figure 1.**
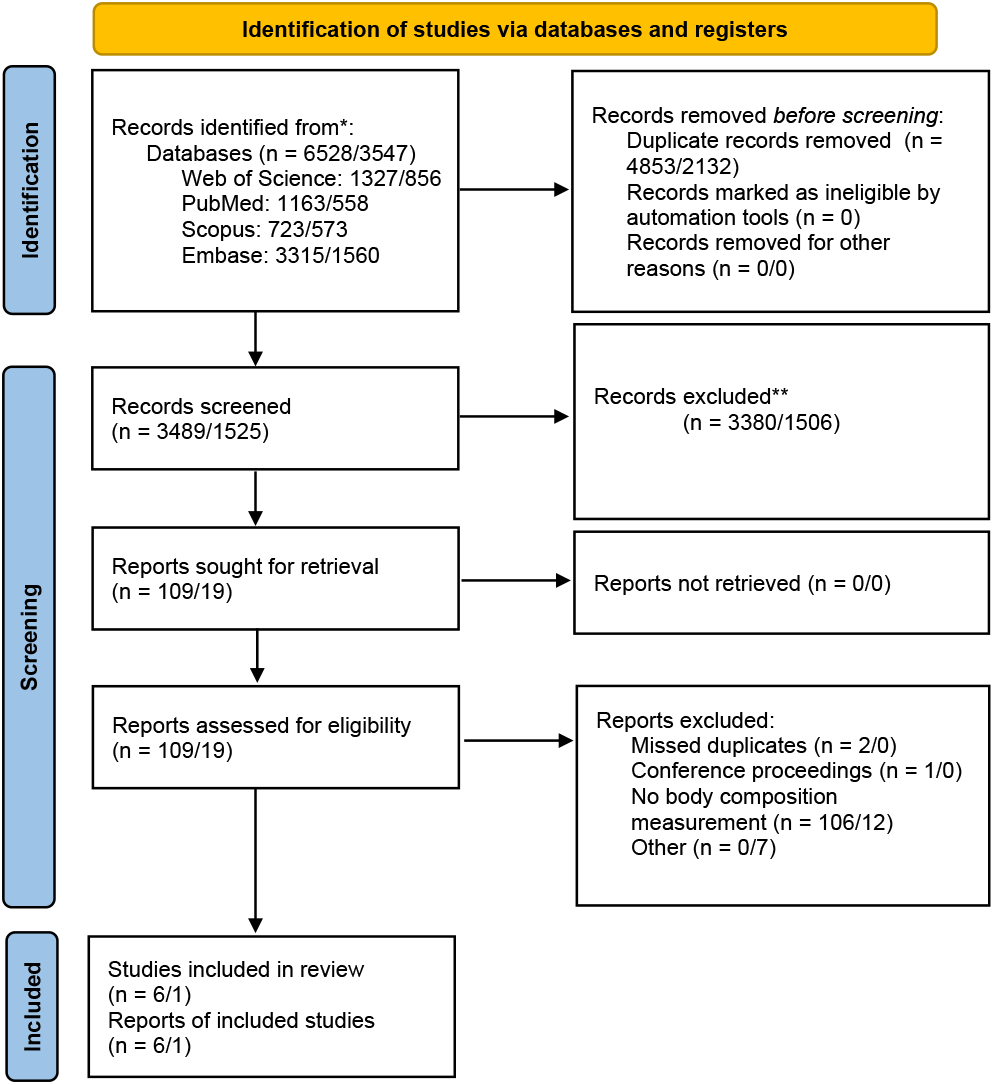
Search strategy. Results are displayed from the initial and updated searches (1^st^ search/2^nd^ search).

### Body mass change

Semaglutide treatment resulted in a significant non-linear body mass loss of 0.04 kg per day (95%CI [0.03-0.05], z = 7.64, p < 0.0001). The significant negative squared time coefficient (−4.6 × 10^-5^ kg, 95%CI [-7.7 × 10^-5^, -1.4 × 10^-5^], z = -2.8, p = 0.004), indicates a slowing in weight loss at longer treatment durations (Figure 2). There was some unexplained variability in the observed effects, with some residual heterogeneity remaining (QE_(df = 15)_ = 25.3, p = 0.06). Between-study variance was estimated at σ^2^ = 1.37, likely due to factors not included here such as patient characteristics (e.g., sex, BMI at baseline, dose, mode of administration).

**Figure 2.**
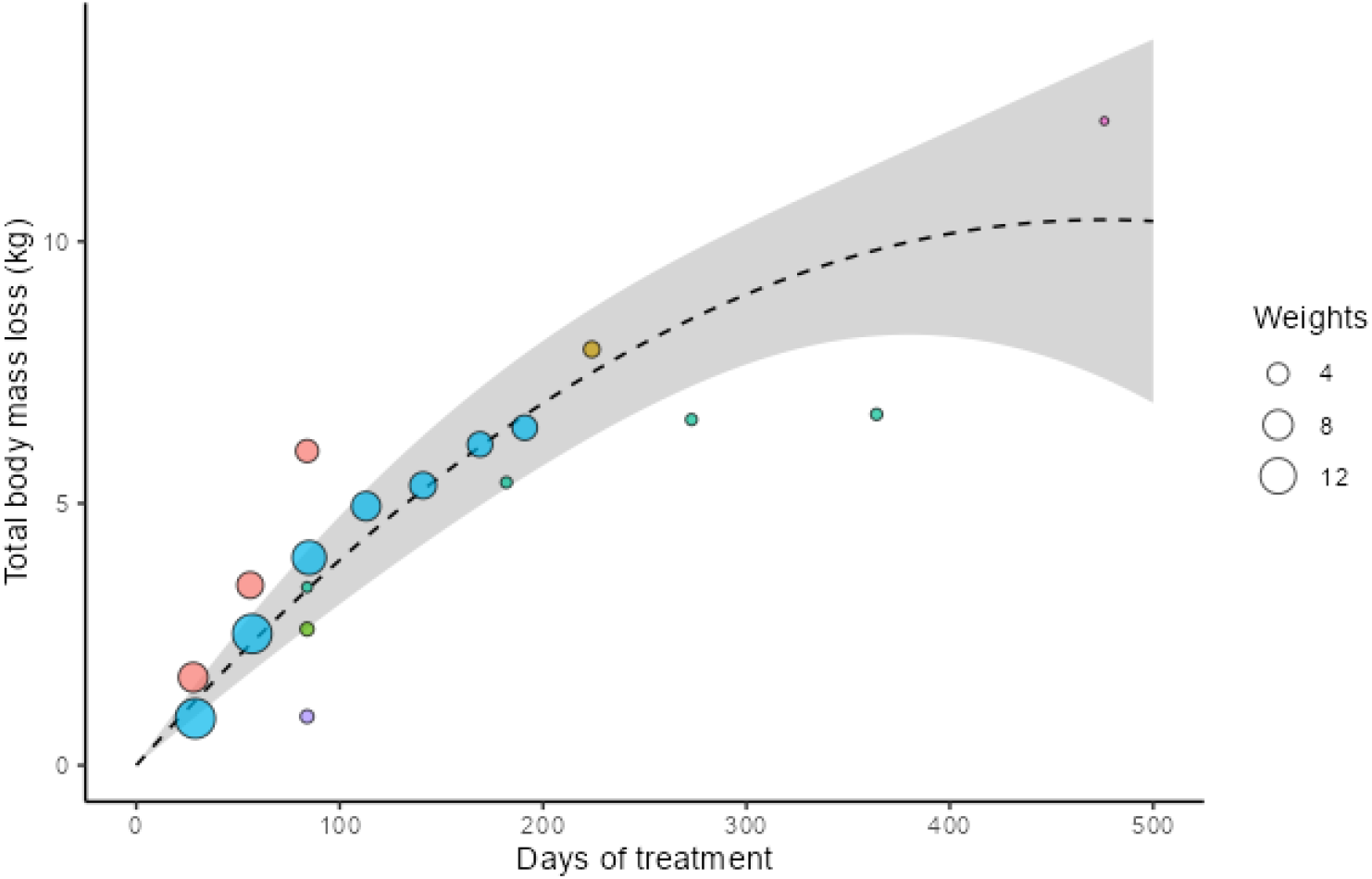
A bubble plot showing the non-linear effects of semaglutide use on body mass. The bubbles represent individual mean differences, and bubble size represent their weights on the analysis. The grey shaded area represents 95% confidence intervals. Bubbles belonging to the same study are shown in the same colour. Red = Blundell et al.^11^; Green = Gibbons et al.^12^; Purple = Ingersen et al.^14^; Blue = Heise et al.^15^; Brown = Eckard et al.^18^; Turqoise = Hansen et al.^17^; Pink = Wilding et al.^23^.

### Fat mass change

A similar pattern to body mass loss was seen for fat mass loss (Figure 3). The estimated effect of semaglutide was 0.03 kg per day (95%CI [0.01, 0.05], z = 2.7, p < 0.007), with a (non-significant) slowing in fat mass loss of -2.8 × 10^-5^ kg per day (95%CI [-8.0 × 10^-5^, 2.5 × 10^-5^], z = -1.03, p = 0.29). With a tau^2^ of 3.68 (SE = 2.96), and an I^2^ of 79.92%, this analysis also presented high heterogeneity.

**Figure 3.**
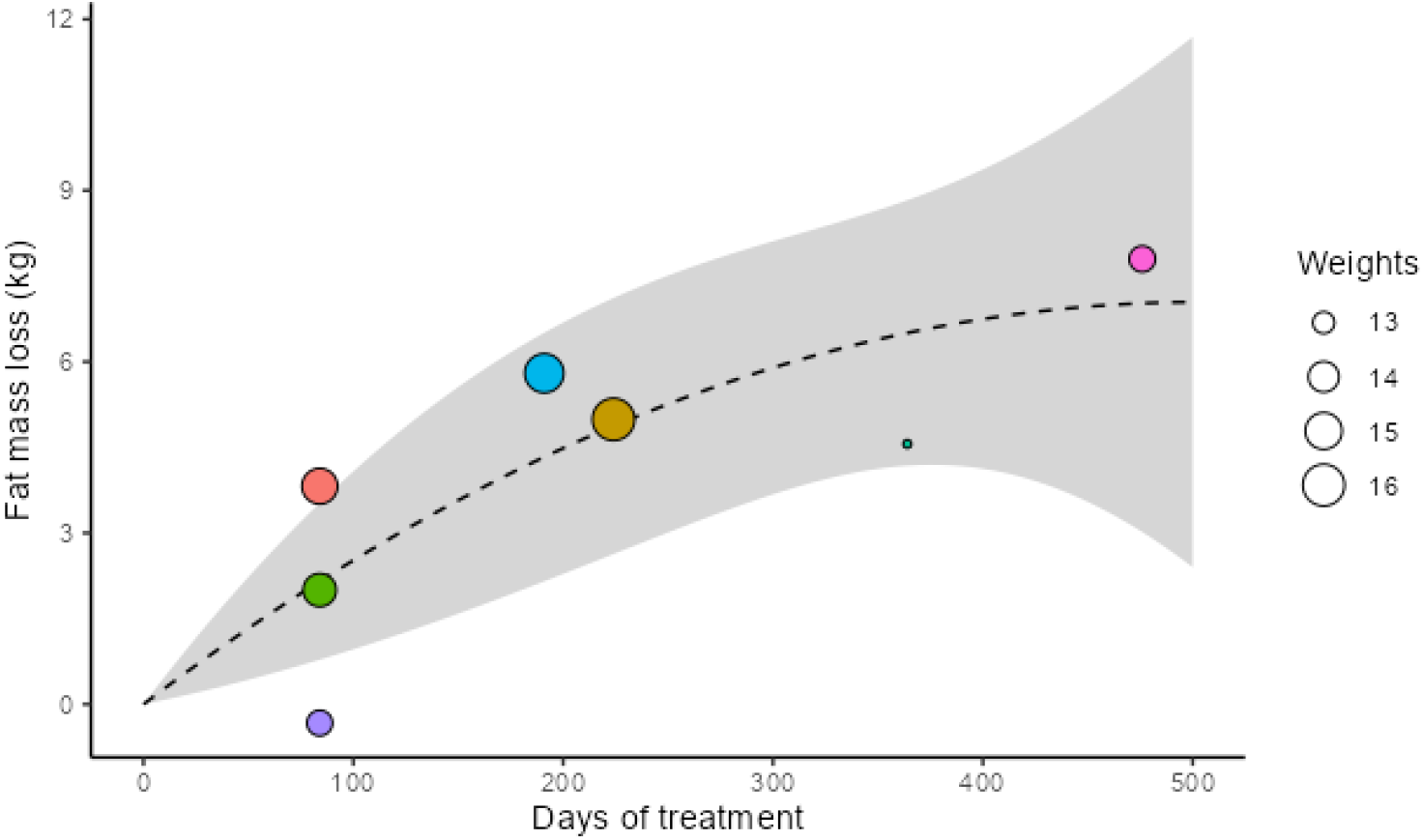
A bubble plot showing effects of semaglutide use on fat-mass over time. The bubbles represent individual mean differences, and bubble size represent their weights on the analysis. The grey shaded area represents 95% confidence intervals. Red = Blundell et al.^11^; Green = Gibbons et al.^12^; Purple = Ingersen et al.^14^; Blue = Heise et al.^15^; Brown = Eckard et al.^18^; Turqoise = Hansen et al.^17^; Pink = Wilding et al.^23^.

### Fat-free mass change

The time effect of semaglutide on fat-free mass loss was defined as linear due to having the best fit, with a reduction of 0.007 kg (95%CI [0.003, 0.010], p = 0.0001, Figure 4) per day. There was substantial residual variability (I^2^ = 82.06%; tau^2^ = 1.10 [SE 0.82]), again suggesting factors not here included might influence outcomes.

**Figure 4.**
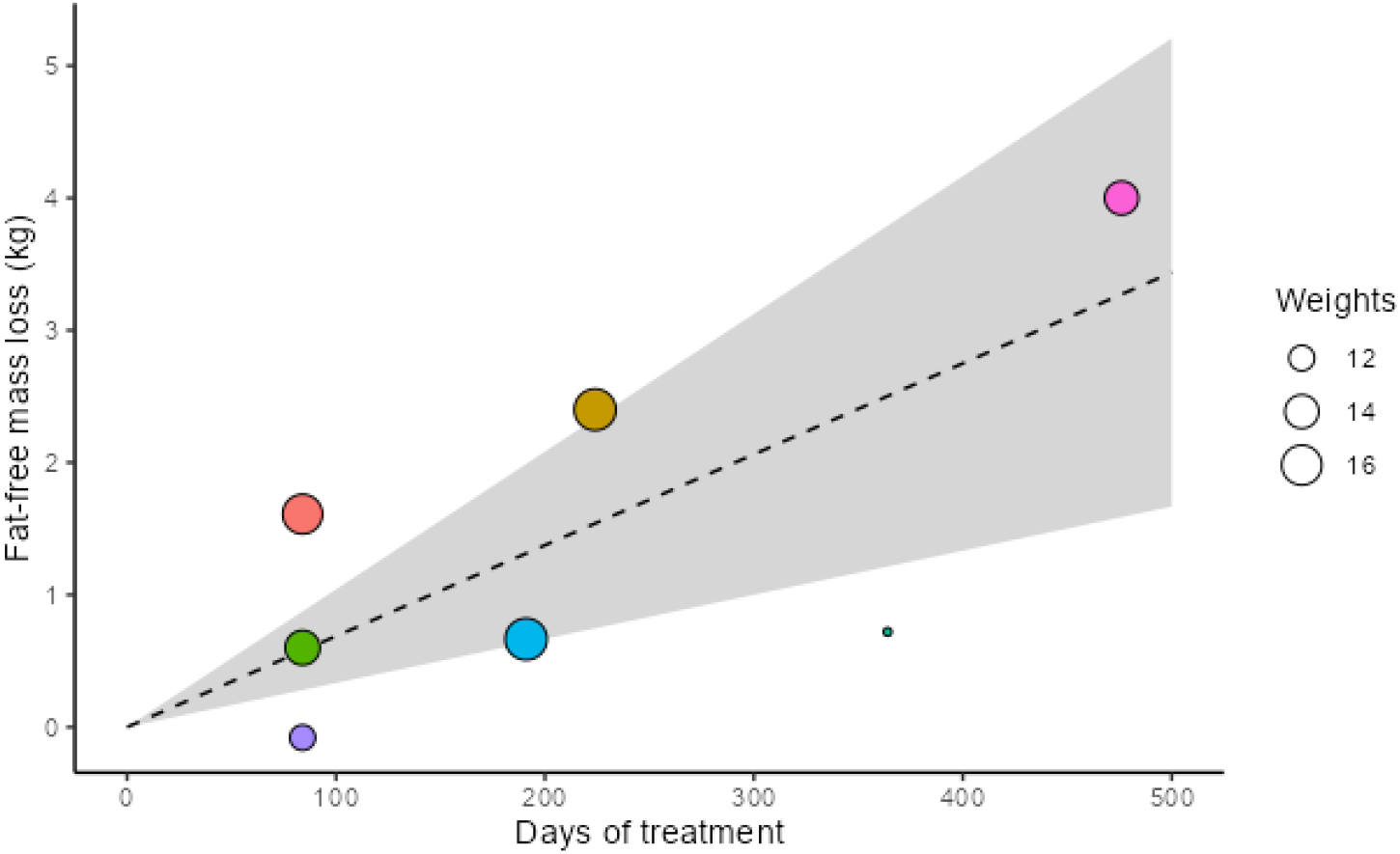
A bubble plot showing effects of semaglutide use on fat-free-mass over time. The bubbles represent individual mean differences, and bubble size represent their weights on the analysis. The grey shaded area represents 95% confidence intervals. Red = Blundell et al.^11^; Green = Gibbons et al.^12^; Purple = Ingersen et al.^14^; Blue = Heise et al.^15^; Brown = Eckard et al.^18^; Turquoise = Hansen et al.^17^; Pink = Wilding et al.^23^.

### Publication bias, risk of bias and GRADE

Some asymmetry was seen in the funnel plots for body mass (Fig. S1), likely due to greater variance in the available time points between studies. For example, Wilding et al.^23^ was the only study with a duration of more than 400 days, with the highest raw residual value observed (2.22) and high variance (2.19), which is expected due to the duration of the intervention. Blundell et al.^11^ showed high raw residual (2.06) on day 86, and a small sample size (13 participants in the semaglutide condition). Both observed higher than expected mean weight loss at the semaglutide condition. On the other hand, the higher variance (∼2.35) and lower than expected weight loss (raw residual = -1.96) in Hansen et al.^17^ after 1 year of treatment resulted in the observed deviations. In Ingersen et al.^14^, participants received exercise training in concomitance with semaglutide, resulting in a comparatively low observed effect (raw residual = -1.56), and relatively high variance (1.62). A sensitivity analysis was performed with the exclusion of these outcomes, resulting in very similar coefficients (0.053 kg/day; 95%CI [0.04, 0.07], p < 0.0001; and -9.3 × 10^-5^ kg/day^2^, 95% CI [-1.6 × 10^-4^, -2.9 × 10^-5^], p = 0.004). The new funnel plot with the exclusion of these outcomes had no evidence of asymmetry (figure not shown) and thus, publication bias was discarded. For fat mass and fat-free mass, no asymmetry was observed, therefore publication bias was discarded. The overall risk of bias was considered low (Fig. S2 in Supporting information). For GRADE (Table S4 in Supporting information), analyses were downgraded due to the high I^2^ values and residual heterogeneity. There were no further downgrades and no upgrades meaning all outcomes had moderate quality.

## Discussion

This systematic review and meta-analysis of seven randomised controlled trials including 452 participants living with obesity showed that semaglutide induces significant body mass reductions over time. The pattern of weight loss was non-linear, with the rate of loss slowing as treatment duration increased. Approximately 75% of total body mass lost was attributable to reductions in fat mass, suggesting that semaglutide preferentially promotes fat reduction. Fat mass losses also exhibited a slowing trend over time, although this did not reach statistical significance. In contrast, fat-free mass declined in a linear fashion, making it difficult to confidently state that the remaining 25% of weight loss is entirely due to fat-free mass. The certainty of the evidence for all outcomes was considered moderate.

Our findings confirm the efficacy of semaglutide to induce weight loss in individuals living with obesity, consistent with prior meta-analyses^6-8,24^. However, the present findings differ from earlier systematic reviews and meta-analyses in several key aspects. We showed that the pattern of body mass losses is consistent with trajectories observed in pharmacological, surgical and lifestyle interventions, where early rapid losses tend to plateau over time^25^. Most of the weight loss with semaglutide appears due to reductions in fat mass, and despite fat-free mass losses, improvement in body composition are shown (i.e., greater lean-to-fat mass ratio)^23^. Importantly, the magnitude of losses in fat-free mass with semaglutide appear more conservative than those following more intensive obesity interventions such as bariatric surgery. Average fat-free mass losses at 3-month (−4.25 kg [95%CI -6.30, -2.20]) and 12-month (−8.23 kg [95%CI -5.73, -10.74]) post-surgery^26^ are well in excess of those estimated at similar time with semaglutide based upon the current data (90-day: -0.63 kg [95% CI -0.27, -0.90]; 12-month: -2.56 kg [95%CI -1.10, -3.65]).

Despite these favourable results with semaglutide, the observed reductions in fat-free mass over time highlight the importance of implementing strategies aimed at preserving fat-free mass, particularly in elderly and sarcopenic individuals^10^. Only one study incorporated exercise training concomitant to semaglutide application^14^, namely 12 weeks of aerobic training, and showed no changes in lean body mass with semaglutide (+0.2 ± 2.7 kg) perhaps representative of the exercise’s ability to protect muscle. However, participants had already taken semaglutide for 20 weeks prior to initiating exercise, with an average -1.6 ± 2.0 kg loss in lean mass. Another study provided semaglutide alongside a lifestyle intervention in which counselling was provided to encourage a daily 500 kcal deficit and increased physical activity^5,23^. Combining pharmacological therapies like semaglutide with resistance exercise^27-29^ and nutritional strategies including increased dietary protein^30,31^ may protect fat-free mass and further enhance body composition outcomes and long-term metabolic health. Such multimodal strategies may optimize long-term outcomes and support healthier weight loss trajectories^32^. From a clinical perspective, practitioners prescribing semaglutide should be encouraged to monitor not only body weight but also changes in body composition over time.

Several observational and non-randomized studies have shown no relevant losses in fat-free mass with semaglutide^33-35^. We chose to include only randomized controlled trials (RCTs) in our meta-analysis. This decision was made to ensure the highest possible standard of evidence for causal inference since RCTs are widely regarded as the gold standard for establishing causality due to their use of random allocation, which minimizes bias and confounding factors^36^. Despite this, high between-study heterogeneity was observed for all body composition outcomes. This variability is likely due to differences in study protocols, participant characteristics (e.g., sex, age, baseline BMI), treatment doses, and administration routes. However, the limited availability of stratified data precluded formal subgroup or meta-regression analyses. Future analyses should consider stratifying by these factors to better understand which populations benefit most and how treatment could be optimized.

### Limitations

One of the primary limitations of this study is the small number of included trials which reduced the overall confidence in the estimates. Our searches identified seven studies totalling 452 participants that measured body composition, though this is only a fraction of the total number of participants since some assessed body composition only in a sub-group^5^. We initially aimed to perform an individual participant data meta-analysis (CRD42024564458; Table S5 in in Supporting information) to overcome limitations related to low trial numbers. However, only two studies made individual participant data available^14,15^. Although most studies stated that anonymized datasets would be shared upon request, ultimately most datasets were not made available. Furthermore, body composition was only assessed at study endpoints, restricting the ability to model changes over time. Most studies measured fat-free or lean body mass, measurements that are comprised of more than muscle (eg, bone, organs, and water), making it difficult to isolate skeletal muscle losses. We urge future studies to assess body composition at multiple timepoints and to adopt transparent data sharing practices to facilitate practical application and robust meta-analytics. Additionally, while several studies employed similar semaglutide dosing strategies (typically providing incremental subcutaneous injections up to 1 mg), one study dosed up to 2.4 mg^5^ while another provided semaglutide orally^12^. Although this limited our ability to investigate a dose-response relationship, it strengthens the conclusions made here based upon the standard therapeutic doses.

## Conclusions

This systematic review and meta-analysis showed that semaglutide significantly reduces body weight in individuals living with obesity, with most of the reduction attributable to fat mass losses. While fat-free mass also declined over time, losses were proportionally smaller and followed a linear trajectory. These findings suggest semaglutide promotes meaningful improvements in body composition. However, the observed reduction in fat-free mass over longer treatment durations underscores the importance of integrating alternate strategies to preserve lean mass. Given the moderate certainty of evidence and high between-study heterogeneity, future randomized trials with detailed body composition assessments and open data sharing practices are warranted to optimize treatment strategies and better understand subgroup responses.

## Data Availability

Data are included in supporting information alongside the manuscript.

## Acknowledgements

The authors are grateful those who provided data upon request. This publication is based on research using data from data contributors Eli Lilly that has been made available through Vivli, Inc (https://vivli.org/). Vivli has not contributed to or approved, and Vivli and Eli Lilly are not in any way responsible for, the contents of this publication.

GG and AB-P are speakers for Novo Nordisk^®^. Novo Nordisk^®^ had no input whatsoever into the current manuscript. BS acknowledges the receipt of a personal research grant from São Paulo Research Foundation (FAPESP 2021/06836-0). The remaining authors report no conflicts of interest related to the content of the current manuscript. No specific funding was received for this work.

