## Supporting information for "Body Composition Changes with Semaglutide: A Systematic Review and Meta-Analysis"

**Table. S1.** Example searches
**Table S2.** Extracted data
**Table S3.** Included studies
**Fig. S2.** Funnel plots
**Fig. S3.** Risk of bias
**Table S4.** GRADE
**Table S5.** Deviations from registered study protocol

**Table S1.** Example searches

Results

(ozempic OR semaglutide) AND (weight OR fat OR muscle OR lean) AND mass

Search > Mapping Date Sources Fields Quick limits EBM Pub. types

**Results Filters** Apply >

Sources
Drugs
Diseases
Devices
Floating Subheadings
Age
Gender
Study types
Publication types
Journal titles
Publication years
Authors
Conference Abstracts
Drug Trade Names
Drug Manufacturers
Device Trade Names
Device Manufacturers

Apply >

**History** Expand

1,135 results for search #5 Set email alert Set RSS feed Search details
Index miner

1 — 25

Select number of items Selected: 0 (clear)Sho

**1** 2024 UPDATE: the Brazilian Diabetes Society position on the management of metabolic dysfunction-associated steatotic liver disease (MASLD) in people with prediabetes or type 2 diabetes
Godoy-Matos A.F., Valério C.M., Silva Júnior W.S., de Araujo-Neto J.M., Bertoluci M.C.
***Diabetology and Metabolic Syndrome*** 2024 **16:1** Article Number 23
Embase NURSING Abstract
Index Terms View Full Text Similar records

**2** Obesity Treatments to Improve Type 1 Diabetes (OTID): a randomized controlled trial of the combination of glucagon-like peptide 1 analogues and sodium-glucose cotransporter 2 inhibitors—protocol for Obesity Treatments to Improve Type 1 Diabetes (the OTID trial)
Al-Ozairi E., Narula K., Miras A.D., Taghadom E., Samad A.E., Al Kandari J., Alyosef A., Mashankar A., Al-Najim W., le Roux C.W.
***Trials*** 2024 **25:1** Article Number 129
Embase MEDLINE Abstract
Index Terms View Full Text

immunosurveillance and increases cancer risk
Piening A., Ebert E., Gottlieb C., Khojandi N., Kuehm L.M., Hoft S.G., Pyles K.D., McCommis K.S., DiPaolo R.J., Ferris S.T., Alspach E., Teague R.M.
***Nature Communications*** 2024 **15:1** Article Number 2835
Embase MEDLINE
Abstract
Index Terms
View Full Text
Similar records

4 Cost-effectiveness of semaglutide 2.4 mg in chronic weight management in Portugal
Silva Miguel L., Soares M., Olivieri A., Sampaio F., Lamotte M., Shukla S., Conde V., Freitas P., Costa J., Borges M.
***Diabetology and Metabolic Syndrome*** 2024 **16:1** Article Number 97
Embase NURSING
Abstract
Index Terms
View Full Text
Similar records

5 Semaglutide ameliorates cardiac remodeling in male mice by optimizing energy substrate utilization through the Creb5/NR4a1 axis
Ma Y.-L., Kong C.-Y., Guo Z., Wang M.-Y., Wang P., Liu F.-Y., Yang D., Yang Z., Tang Q.-Z.
***Nature Communications*** 2024 **15:1** Article Number 4757
Embase MEDLINE
Abstract
Index Terms
View Full Text
Similar records

6 Current perspectives in obesity management: unraveling the impact of different therapy approach in real life obesity care
Khorrami Chokami K., Khorrami Chokami A., Cammarata G., Piras G., Albertelli M., Gatto F., Vera L., Ferone D., Boschetti M.
***Journal of Translational Medicine*** 2024 **22:1** Article
Embase MEDLINE NURSING
Abstract
Index Terms
View Full Text
Similar records

7 Discovery of novel glucagon-like peptide 1/cholecystokinin 1 receptor dual agonists
Chenxu Z., Lidan S., Guoqiang H.,

Index Terms View Full Text Similar records

8
Semaglutide and health risk: Development and validation of a LC-HRMS method for testing semaglutide in whole blood and application to real cases
Arbouche N., Blanchot A., Raul J.S., Kintz P.
***Journal of Chromatography B: Analytical Technologies in the Biomedical and Life Sciences*** 2024 **1242** Article Number 124187
Embase MEDLINE
Abstract Index Terms View Full Text Similar records

9
Semaglutide and NT-proBNP in Obesity-Related HFpEF: Insights From the STEP-HFpEF Program
Petrie M.C., Borlaug B.A., Butler J., Davies M.J., Kitzman D.W., Shah S.J., Verma S., Jensen T.J., Einfeldt M.N., Liisberg K., Perna E., Sharma K., Ezekowitz J.A., Fu M., Melenovský V., Ito H., Lelonek M., Kosiborod M.N.
***Journal of the American College*** [illegible] **:1** (27-40)
Embase MEDLINE NURSING
Abstract Index Terms
View Full Text Similar records

10
A commentary review on endoscopic sleeve gastroplasty: Indications, outcomes and future implications
Abuawwad M., Tibude A., Bansi D., Idris I., Madhok B.
***Diabetes, Obesity and Metabolism*** 2024 **26:7** (2546-
Embase MEDLINE NURSING
Abstract Index Terms
View Full Text Similar records

11
Factors associated with treatment responses to pioglitazone in patients with steatotic liver disease: A 3-year prospective cohort study
Chang M.-L., Tai J., Cheng J.-S., Chen W.-T., Yang S.-S., Chiu C.-H., Chien R.-N.
***Diabetes, Obesity and***
Embase MEDLINE NURSING

12 Identification and utility exploration of a highly potent and long-acting bullfrog GLP-1 analogue in GLP-1 and amylin combination therapy

Sun X., Yang D., Li Y., Shi J., Zhang X., Yi T.

***Peptides*** 2024 **177** Article Number 171203

Embase MEDLINE

Abstract Index Terms View Full Text Similar records

13 Weight gain during midlife: Does race/ethnicity influence risk?

Persons P.A., Williams L., Fields H., Mishra S., Mehta R.

***Maturitas*** 2024 **185** Article Number [illegible]

Embase MEDLINE NURSING

Abstract Index Terms View Full Text Similar records

14 Long-Term Efficacy and Safety of Once-Weekly Semaglutide for Weight Loss in Patients Without Diabetes: A Systematic Review and Meta-Analysis of Randomized Controlled Trials

Moiz A., Levett J.Y., Filion K.B., Peri K., Reynier P., Eisenberg M.J.

***American Journal of*** [illegible] 121-130)

Embase MEDLINE NURSING

Abstract Index Terms View Full Text Similar records

15 GLP-1 receptor agonists for weight reduction in people living with obesity but without diabetes: a living benefit–harm modelling study

Moll H., Frey E., Gerber P., Geidl B., Kaufmann M., Braun J., Beuschlein F., Puhan M.A., Yebyo H.G.

***eClinicalMedicine*** 2024 **73** Article Number 1[illegible]

Embase

Abstract Index Terms View Full Text Similar records

16 Tirzepatide Improved Markers of Islet Cell Function and Insulin Sensitivity in People With T2D (SURPASS-2)

Frias J.P., De Block C., Brown K.,

1753)
Embase MEDLINE
Abstract
Index Terms
View Full Text
Similar records

17
Approach to the Patient With Type 2 Diabetes Requiring Add-On Medication
Baum H.B.A.
***Journal of Clinical Endocrinology and Metabolism*** 2024 **109:7** (e1506-e1512)
Embase MEDLINE
Abstract
Index Terms
View Full Text
Similar records

18
Treatment of Obesity in Heart Failure: The New Era in the Aftermath of the STEP-HFpEF and SELECT Trials
Bozkurt B.
***JACC: Heart Failure*** 2024 **12:7** (1309-1313)
Embase NURSING
[No abstract available]
Index Terms
View Full Text
Similar records

19
Semaglutide mitigates testicular damage in diabetes by inhibiting ferroptosis
Zhou L., Dong M., Feng G., Zhang Y., Wang J., Kang H., Dong Z., Ning J., Zhao Z., Wang C.
***Biochemical and Biophysical Research Communications*** 2024 **715** Article Number 149996
Embase MEDLINE
Abstract
Index Terms
View Full Text
Similar records

20
Effects of Dapagliflozin on Body Composition and Its Relation to Hemodynamics in Heart Failure With Preserved Ejection Fraction
Naser J.A., Tada A., Harada T., Reddy Y.N.V., Carter R.E., Testani J.M., Jensen M.D., Borlaug B.A.
***Circulation*** 2024 **149:25** (2026-2028)
Embase MEDLINE NURSING
[No abstract available]
Index Terms
View Full Text
Similar records

21
Real-world clinical evidence of oral semaglutide on metabolic abnormalities in subjects with type 2 diabetes: a multicenter retrospective observational

Kurihara H., Aoki S., Miya A., Kameda H., Nakamura A., Atsumi T.
***Endocrine Journal*** 20[illegible]
MEDLINE Abstract Index Terms
View Full Text Similar records

22

Efficacy and Safety of Once-Weekly Subcutaneous Semaglutide in Overweight or Obese Adults: A Systematic Review with Meta-Analysis
Dorneles G., Algeri E., Lauterbach G., Pereira M., Fernandes B.
***Experimental and Clinical Endocrinology and Diabetes*** 2024 **132** [illegible]7)
Embase MEDLINE Abstract
Index Terms View Full Text Similar records

23

Favorable liver and skeletal muscle changes in patients with MASLD and T2DM receiving glucagon-like peptide-1 receptor agonist: A prospective cohort study
Kakegawa T., Sugimoto K., Saito K., Yunaiyama D., Araki Y., Wada T., Takahashi H., Yoshimasu Y., Takeuchi H., Itoi T.
***Medicine (United States)*** 2024 **103:23** Article Number e38444
Embase MEDLINE Abstract
Index Terms View Full Text Similar records

24

Effectiveness of oral semaglutide on glucose control and body weight up to 18 months: a multicenter retrospective real-world study
Bonora B.M., Russo G., Leonetti F., Strazzabosco M., Nollino L., Aimaretti G., Giaccari A., Broglio F., Consoli A., Avogaro A., Fadini G.P.
***Journal of Endocrinological Investigation*** 2024 **47:6** (1395-1403)
Embase MEDLINE Abstract
Index Terms View Full Text Similar records

25

Case report of the successful use of semaglutide to achieve target BMI prior to renal transplant in two patients with end-stage-kidney-

Index Terms

View Full Text
Similar records

Records per page 25



Copyright © 2024 Elsevier Limited except certain content provided by third parties. Embase is a trademark of Elsevier Limited. All rights are reserved, including those for text and data mining, AI training, and similar technologies.
We use cookies to help provide and enhance our service and tailor content. Cookie Settings

An official website of the United States government
Here's how you know



1 Effect of Weekly Subcutaneous **Semaglutide** vs Daily Liraglutide on **Body Weight** in Adults With Overweight or Obesity Without Diabetes: The STEP 8 Randomized Clinical Trial.
Rubino DM, Greenway FL, Khalid U, O'Neil PM, Rosenstock J, Sørrig R, Wadden TA, Wizert A, Garvey WT; STEP 8 Investigators.
JAMA. 2022 Jan 11;327(2):138-150. doi: 10.1001/jama.2021.23619.
PMID: 35015037 Free PMC article. Clinical Trial.
View PDF

2 Effect of Continued Weekly Subcutaneous **Semaglutide** vs Placebo on **Weight** Loss Maintenance in Adults With Overweight or Obesity: The STEP 4 Randomized Clinical Trial.
Rubino D, Abrahamsson N, Davies M, Hesse D, Greenway FL, Jensen C, Lingvay I, Mosenzon O, Rosenstock J, Rubio MA, Rudofsky G, Tadayon S, Wadden TA, Dicker D; STEP 4 Investigators.
JAMA. 2021 Apr 13;325(14):1414-1425. doi: 10.1001/jama.2021.3224.
PMID: 33755728 Free PMC article. Clinical Trial.
View PDF

3 Once-Weekly **Semaglutide** in Adults with Overweight or Obesity.
Wilding JPH, Batterham RL, Calanna S, Davies M, Van Gaal LF, Lingvay I, McGowan BM, Rosenstock J, Tran MTD, Wadden TA, Wharton S, Yokote K, Zeuthen N, Kushner RF; STEP 1 Study Group.
N Engl J Med. 2021 Mar 18;384(11):989-1002. doi: 10.1056/NEJMoa2032183. Epub 2021 Feb 10.
PMID: 33567185 Clinical Trial.
View PDF

4 **Weight** regain and cardiometabolic effects after withdrawal of **semaglutide**: The STEP 1 trial extension.
Wilding JPH, Batterham RL, Davies M, Van Gaal LF, Kandler K, Konakli K, Lingvay I, McGowan BM, Oral TK, Rosenstock J, Wadden TA, Wharton S, Yokote K, Kushner RF; STEP 1 Study Group.
Diabetes Obes Metab. 2022 Aug;24(8):1553-1564. doi: 10.1111/dom.14725. Epub 2022 May 19.
PMID: 35441470 Free PMC article. Clinical Trial.

5 **Semaglutide** 2.4 mg once a week in adults with overweight or obesity, and type 2 diabetes (STEP 2): a randomised, double-blind, double-dummy, placebo-controlled, phase 3 trial.
Davies M, Færch L, Jeppesen OK, Pakseresht A, Pedersen SD, Perreault L, Rosenstock J, Shimomura I, Viljoen A, Wadden TA, Lingvay I; STEP 2 Study Group.
Lancet. 2021 Mar 13;397(10278):971-984. doi: 10.1016/S0140-6736(21)00213-0. Epub 2021 Mar 2.
PMID: 33667417 Clinical Trial.

6 **Semaglutide** in Patients with Heart Failure with Preserved Ejection Fraction and Obesity.
Kosiborod MN, Abildstrøm SZ, Borlaug BA, Butler J, Rasmussen S, Davies M, Hovingh GK, Kitzman DW, Lindegaard ML, Møller DV, Shah SJ, Treppendahl MB, Verma S, Abhayaratna W, Ahmed FZ, Chopra V, Ezekowitz J, Fu M, Ito H, Lelonek M, Melenovsky V, Merkely B, Núñez J, Perna E, Schou M, Senni M, Sharma K, Van der Meer P, von Lewinski D, Wolf D, Petrie MC; STEP-HFpEF Trial Committees and Investigators.
N Engl J Med. 2023 Sep 21;389(12):1069-1084. doi: 10.1056/NEJMoa2306963. Epub 2023 Aug 25.
PMID: 37622681 Clinical Trial.
View PDF

7 Safety of **Semaglutide**.
Smits MM, Van Raalte DH.
Front Endocrinol (Lausanne). 2021 Jul 7;12:645563. doi: 10.3389/fendo.2021.645563. eCollection 2021.
PMID: 34305810 Free PMC article. Review.
View PDF

8 Once-Weekly **Semaglutide** in Adolescents with Obesity.
Weghuber D, Barrett T, Barrientos-Pérez M, Gies I, Hesse D, Jeppesen OK, Kelly AS, Mastrandrea LD, Sørrig R, Arslanian S; STEP TEENS Investigators.
N Engl J Med. 2022 Dec 15;387(24):2245-2257. doi: 10.1056/NEJMoa2208601. Epub 2022 Nov 2.
PMID: 36322838 Free PMC article. Clinical Trial.

9 GLP-1 receptor agonists in the treatment of type 2 diabetes - state-of-the-art.
Nauck MA, Quast DR, Wefers J, Meier JJ.
Mol Metab. 2021 Apr;46:101102. doi: 10.1016/j.molmet.2020.101102. Epub 2020 Oct 14.
PMID: 33068776 Free PMC article. Review.

10 **Semaglutide** for the treatment of obesity.
Chao AM, Tronieri JS, Amaro A, Wadden TA.
Trends Cardiovasc Med. 2023 Apr;33(3):159-166. doi: 10.1016/j.tcm.2021.12.008. Epub 2021 Dec 21.
PMID: 34942372 Free PMC article. Review.



FOLLOW NCBI

Connect with NLM

National Library of Medicine
8600 Rockville Pike
Bethesda, MD 20894

Web Policies
FOIA
HHS Vulnerability Disclosure

Help
Accessibility
Careers

NLM NIH HHS USA.gov

ABCD USP .periodicos. CAPES

Scopus

Welcome to a more intuitive and efficient search experience. See what is new

Save search

Set search alert

Advanced query

Search within
Article title, Abstract, Keywords

Search documents *
( ozempic OR semaglutide ) AND ( weight OR fat OR muscle OR lean AND mass )

+ Add search field

Reset Search

Documents | Preprints (Beta) | Patents | Secondary documents | Research data ↗

### 723 documents found

Analyze results ↗

#### Refine search

Search within results

#### Filters

**Year**

Range Individual

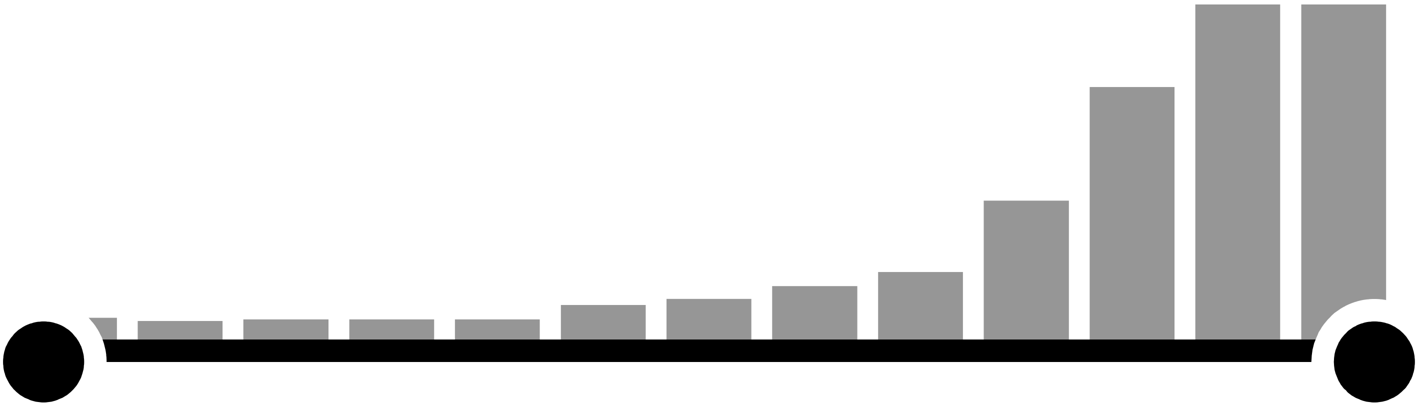

from – to

**Subject area**

| | |
|---|---|
| Medicine | 631 |
| Biochemistry, Genetics and Molecular Biology | 199 |
| Pharmacology, Toxicology and Pharmaceutics | 71 |
| Nursing | 64 |
| Health Professions | 15 |

Show all

**Document type**

| | |
|---|---|
| Article | 407 |
| Review | 201 |

| | |
|---|---|
| Note | 48 |
| Editorial | 29 |
| Letter | 28 |

Show all

**Language**

| | |
|---|---|
| English | 716 |
| French | 4 |
| Spanish | 3 |
| Chinese | 2 |
| German | 1 |

Show all

**Keyword**

| | |
|---|---|
| Semaglutide | 701 |
| Human | 669 |
| Body Mass | 587 |
| Obesity | 507 |
| Body Weight Loss | 452 |

Show all

**Country/territory**

**Source type**

**Source title**

**Author name**

**Publication stage**

**Affiliation**

**Funding sponsor**

**Open access**

Export filter counts

All Export Download Citation overview ••• More Show all abstracts Sort by Date (newest)

| | Document title | Authors | Source | Year | Citations |
|---|---|---|---|---|---|
| 1 | Article • *Open access*<br>**Current perspectives in obesity management: unraveling the impact of different therapy approach in real life obesity care**<br>Show abstract Buscar na USP View at Publisher Related documents | Khorrami Chokami, K., Khorrami Chokami, A., Cammarata, G., ... Ferone, D., Boschetti, M. | Journal of Translational Medicine , 22(1), 536 | 2024 | 0 |
| 2 | Article • *Open access*<br>**Semaglutide ameliorates cardiac remodeling in male mice by optimizing energy substrate utilization through the Creb5/NR4a1 axis**<br>Show abstract Buscar na USP View at Publisher Related documents | Ma, Y.-L., Kong, C.-Y., Guo, Z., ...Yang, Z., Tang, Q.-Z. | Nature Communications , 15(1), 4757 | 2024 | 0 |

Article • *Open access*

| | Document title | Authors | Source | Year | Citations |
|---|---|---|---|---|---|
| 3 | **Cost-effectiveness of semaglutide 2.4 mg in chronic weight management in Portugal**<br>Show abstract Buscar na USP View at Publisher Related documents | Silva Miguel, L., Soares, M., Olivieri, A., ... Costa, J., Borges, M. | Diabetology and Metabolic Syndrome , 16(1), 97 | 2024 | 0 |

**Discover early research ideas**

View preprints published by authors to have an early idea of upcoming research documents.

View 9 preprints

| | Document title | Authors | Source | Year | Citations |
|---|---|---|---|---|---|
| 4 | Article • *Open access*<br>**Obesity-related T cell dysfunction impairs immunosurveillance and increases cancer risk**<br>Show abstract Buscar na USP View at Publisher Related documents | Piening, A., Ebert, E., Gottlieb, C., ...Alspach, E., Teague, R.M. | Nature Communications , 15(1), 2835 | 2024 | 0 |
| 5 | Article • *Open access*<br>**Obesity Treatments to Improve Type 1 Diabetes (OTID): a randomized controlled trial of the combination of glucagon-like peptide 1 analogues and sodium-glucose cotransporter 2 inhibitors—protocol for Obesity Treatments to Improve Type 1 Diabetes (the OTID trial)**<br>Show abstract Buscar na USP View at Publisher Related documents | Al-Ozairi, E., Narula, K., Miras, A.D., ...Al-Najim, W. , le Roux, C.W. | Trials, 25(1), 129 | 2024 | 0 |
| 6 | Review • *Open access*<br>**2024 UPDATE: the Brazilian Diabetes Society position on the management of metabolic dysfunction-associated steatotic liver disease (MASLD) in people with prediabetes or type 2 diabetes**<br>Show abstract Buscar na USP View at Publisher Related documents | Godoy-Matos, A.F., Valério, C.M., Silva Júnior, W.S., de Araujo-Neto, J.M., Bertoluci, M.C. | Diabetology and Metabolic Syndrome , 16(1), 23 | 2024 | 2 |
| 7 | Review<br>**Rare cutaneous adverse reactions associated with GLP-1 agonists: a review of the published literature**<br>Show abstract Buscar na USP View at Publisher Related documents | Salazar, C.E., Patil, M.K., Aihie, O., Cruz, N., Nambudiri, V.E. | Archives of Dermatological Research , 316(6), 248 | 2024 | 0 |
| 8 | Article<br>**Discovery of novel glucagon-like peptide 1/cholecystokinin 1 receptor dual agonists**<br>Show abstract Buscar na USP View at Publisher Related documents | Chenxu, Z., Lidan, S., Guoqiang, H., ...Xiaoyi, S., Qian, L. | European Journal of Pharmaceutical Sciences , 199, 106818 | 2024 | 0 |
| 9 | Article<br>**Semaglutide and health risk: Development and validation of a LC-HRMS method for testing semaglutide in whole blood and application to real cases**<br>Show abstract Buscar na USP View at Publisher Related documents | Arbouche, N., Blanchot, A. , Raul, J.S., Kintz, P. | Journal of Chromatography B: Analytical Technologies in the Biomedical and Life Sciences , 1242, 124187 | 2024 | 0 |
| 10 | Article • *Open access*<br>**Semaglutide and NT-proBNP in Obesity-Related HFpEF: Insights From the STEP-HFpEF Program** | Petrie, M.C., Borlaug, B.A. , Butler, J., ...Lelonek, M., Kosiborod, M.N. | Journal of the American College of Cardiology , 84(1), pp. 27–40 | 2024 | 1 |

| Document title | Authors | Source | Year | Citations |
| --- | --- | --- | --- | --- |

Show abstract Buscar na USP View at Publisher Related documents

### About Scopus

What is Scopus
Content coverage
Scopus blog
Scopus API
Privacy matters

### Language

日本語版を表示する
查看简体中文版本
查看繁體中文版本
Просмотр версии на русском языке

### Customer Service

Help
Tutorials
Contact us

ELSEVIER

Terms and conditions ↗ Privacy policy ↗

All content on this site: Copyright © 2024 Elsevier B.V. ↗, its licensors, and contributors. All rights are reserved, including those for text and data mining, AI training, and similar technologies. For all open access content, the Creative Commons licensing terms apply.
We use cookies to help provide and enhance our service and tailor content.By continuing, you agree to the use of cookies ↗.

RELX™

**1,327 results from Web of Science Core Collection for:**

Analyze Results | Citation Report | Create Alert

**(Ozempic OR Semaglutide) AND (Weight OR Fat OR Muscle OR Lean mass)** (All Fields)

Search

Did you mean? **(olympic OR Semaglutide) AND (Weight OR Fat OR Muscle OR Lean mass)** (All Fields) | 3,286 results

Add Keywords | **Quick add keywords:** + SEMAGLUTIDE + ORAL SEMAGLUTIDE + TIRZEPA

Publications | You may also like... | Copy query link

### Refine results

Search within results...

#### Quick Filters

| | |
|---|---|
| Highly Cited Papers | 71 |
| Hot Papers | 7 |
| Review Article | 392 |
| Early Access | 69 |
| Open Access | 765 |
| Enriched Cited References | 384 |
| Open publisher-invited reviews | 68 |

#### Publication Years

Show Final Publication Year

| | |
|---|---|
| 2024 | 260 |
| 2023 | 406 |
| 2022 | 238 |
| 2021 | 163 |
| 2020 | 78 |

See all >

#### Document Types

| | |
|---|---|
| Article | 695 |
| Review Article | 392 |
| Meeting Abstract | 137 |
| Editorial Material | 70 |
| Early Access | 69 |

0/1,327 Add To Marked List Export

Sort by Relevance Previous page Next page

1 **Implications of Ozempic and Other Semaglutide Medications for Facial Plastic Surgeons**

Humphrey, CD and Lawrence, AC

Dec 2023 | FACIAL PLASTIC SURGERY 39 (06) , pp.719-721

Obesity is a growing global health concern, leading to various health issues, including diabetes. Semagl ... Show more

Free Full Text From Publisher

5 Citations

7 References

Related records

2 **A systematic review of the effect of semaglutide on lean mass: insights from clinical trials**

Bikou, A; Dermiki-Gkana, F; (...); Kontogiorgis, C

Mar 23 2024 | EXPERT OPINION ON PHARMACOTHERAPY 25 (5) , pp.611-619

IntroductionSemaglutide, a glucagon-like peptide-1 receptor agonist, is associated with significant w ... Show more

View full text

53 References

Related records

12

See all >

Researcher Profiles

Show Researcher Profiles

- Lingvay, Ildiko 41
- Davies, Michael 38
- Rosenstock, Julio 28
- Le Roux, Carel W. 23
- Aroda, Vanita R. 21

See all >

Web of Science Categories

Citation Topics Meso

Citation Topics Micro

Web of Science Index

Affiliations

Affiliation with Department

Publication Titles

Languages

Countries/Regions

Publishers

Research Areas

Open Access

Filter by Marked List

Funding Agencies

Conference Titles

Group Authors

Book Series Titles

Editors

Editorial Notices

3 Oral Semaglutide Induces Loss of Body Fat Mass Without Affecting Muscle Mass in Patients With Type 2 Diabetes

Uchiyama, S; Sada, Y; (...); Tanaka, Y
Jul 2023
|
JOURNAL OF CLINICAL MEDICINE RESEARCH-CANADA
15 (7) , pp.377-383

Enriched Cited References

Background: Excessive body fat may be a major cause of insulin resistance and diabetes. But boc ... Show more

Free Full Text from Publisher

3 Citations
28 References
Related records

4 Clinical effectiveness of semaglutide on weight loss, body composition, and muscle strength in Chinese adults

Xiang, J; Ding, XY; (...); Liang, YZ
Oct 2023
|
EUROPEAN REVIEW FOR MEDICAL AND PHARMACOLOGICAL SCIENCES
27 (20) , pp.9908-9915

OBJECTIVE: The aim of this study was to investigate the clinical effectiveness of semaglutide on w ... Show more

4 Citations
36 References
Related records

5 Oral semaglutide improves body composition and preserves lean mass in patients with type 2 diabetes: a 26-week prospective real-life study

Volpe, S; Lisco, G; (...); Piazzolla, G
Sep 13 2023
| FRONTIERS IN ENDOCRINOLOGY 14

Enriched Cited References

BackgroundOral semaglutide is the first glucagon-like peptide-1 receptor agonist (GLP-1RA) design ... Show more

Free Full Text from Publisher

8 Citations
56 References
Related records

6 As Ozempic's Popularity Soars, Here's What to Know About Semaglutide and Weight Loss

5 Citations

Sustainable Development Goals

For more options, use **Analyze Results**

Suran, M
May 16 2023
|
JAMA-JOURNAL OF THE AMERICAN MEDICAL ASSOCIATION
329 (19) , pp.1627-1629

This Medical News article discusses chronic weight management with semaglutide, sold under the brand names Ozempic and

Full Text at Publisher •••

0
References

7 Use of Dulaglutide, Semaglutide, and Tirzepatide in Diabetes and Weight Management

Powell, J and Taylor, J

Mar 2024 | CLINICAL THERAPEUTICS 46 (3) , pp.289-292

Purpose: Glucagon-like peptide 1 receptor agonists (GLP1-RA) are effective therapies in lowering glycosyl ... Show more

Full Text at Publisher •••

29
References

Related records

8 Semaglutide (Ozempic ® ) Use in Denmark 2018 Through 2023-User Trends and off-Label Prescribing for Weight Loss

Mailhac, A; Pedersen, L; (...); Thomsen, RW

2024 | CLINICAL EPIDEMIOLOGY 16 , pp.307-318

Purpose: A surge in the use of semaglutide injection (Ozempic (R) ) approved to treat type 2 diabetes (1 ... Show more

Free Full Text from Publisher •••

43
References

Related records

9 Efficacy and Safety of Semaglutide in Weight Loss Non-diabetic People

Song, CE; Wang, Y; (...); Wu, HY
May 2024 (Early Access)
|
ENDOCRINE METABOLIC & IMMUNE DISORDERS-DRUG TARGETS

Objective The study aimed to investigate the efficacy and safety of semaglutide in weight loss in non-diabe ... Show more

View full text •••

36
References

Related records

10 Transforming body composition with semaglutide in adults with

12

obesity and type 2 diabetes mellitus

Jiménez, BR; Gómez, PRD; (...); Martínez-Brocca, MA

Jun 4 2024

| FRONTIERS IN ENDOCRINOLOGY 15

Enriched Cited References

Background: Glucagon-like peptide-1 receptor-agonists (GLP-1ra), such as semaglutide, hav ... Show more

Free Full Text from Publisher •••

62 References

Related records

11 GLP-1RA Liraglutide and Semaglutide Improves Obesity-Induced Muscle Atrophy via SIRT1 Pathway

Xiang, J; Qin, LY; (...); Liang, YZ

2023

|

DIABETES METABOLIC SYNDROME AND OBESITY

16 , pp.2433-2446

Background: Obesity is related to the loss of skeletal muscle mass and function (sarcopenia). The ... Show more

Free Full Text from Publisher •••

6 Citations

59 References

Related records

12 Semaglutide for Weight Loss: Was It Worth the Weight?

Novograd, J; Mullally, J and Frishman, WH

Nov-dec 2022 | CARDIOLOGY IN REVIEW

30 (6) , pp.324-329

Obesity is a major public health issue with an increasing prevalence worldwide. Excess body fat is associ ... Show more

Full Text at Publisher •••

5 Citations

48 References

Related records

13 Anti-fatness in the Ozempic era: state of the landscape and considerations for future research

Oswald, F

Jan 2024 (Early Access)

|

FAT STUDIES-AN INTERDISCIPLINARY JOURNAL OF BODY WEIGHT AND SOCIETY

Enriched Cited References

The recent popularity of a new type of drug - semaglutide, brand named Ozempic or Wegovy - for fat lo ... Show more

1 Citation

31 References

View full text •••

Related records

14 Semaglutide alleviates gut microbiota dysbiosis induced by a high-fat diet

Duan, XH; Zhang, L; (...); Qiu, JF
Apr 15 2024
|
EUROPEAN JOURNAL OF PHARMACOLOGY
969

This study investigated the effects of semaglutide (Sema) on the gut microbiota of obese mice induc ... Show more

Full Text at Publisher •••

60 References

Related records

15 Semaglutide's slimming properties shifts the scales towards scarcity and shams

Vermaak, I
2024 | SA PHARMACEUTICAL JOURNAL
91 (1) , pp.74-77

Semaglutide, a glucagon-like peptide-1 receptor agonist (GLP-1 RA), has gained attention for its c ... Show more

•••

19 References

Related records

16 Semaglutide modulates prothrombotic and atherosclerotic mechanisms, associated with epicardial fat, neutrophils and endothelial cells network

García-Vega, D; Sánchez-López, D; (...); Eiras, S
Jan 3 2024
| CARDIOVASCULAR DIABETOLOGY 23 (1)

Enriched Cited References

BackgroundObesity has increased in recent years with consequences on diabetes and other comorbidit ... Show more

Free Full Text from Publisher •••

6 Citations

70 References

Related records

17 Beyond appetite regulation: Targeting energy expenditure, fat oxidation, and lean mass preservation for sustainable weight loss

27 Citations

162 References 12

Christoffersen, BO; Sanchez-Delgado, G; (...); Ravussin, E

Apr 2022 | OBESITY 30 (4) , pp.841-857

New appetite-regulating antiobesity treatments such as semaglutide and agents under investigati ... Show more

Free Published Article From Repository

View full text

Related records

18 Antibody blockade of activin type II receptors preserves skeletal muscle mass and enhances fat loss during GLP-1 receptor agonism

Nunn, E; Jaiswal, N; (...); Titchenell, PM

Feb 2024 | MOLECULAR METABOLISM 80

Objective: Glucagon-like peptide 1 (GLP-1) receptor agonists reduce food intake, producing remarl ... Show more

Free Full Text from Publisher

2 Citations

41 References

Related records

19 Semaglutide reduces fat accumulation in the tongue: A randomized single-blind, pilot study

Jensterle, M; Ferjan, S; (...); Janez, A

Aug 2021 | DIABETES RESEARCH AND CLINICAL PRACTICE 178

Enriched Cited References

Aim: We evaluated the effect of the latest GLP-1 RA semaglutide on tongue fat storage in obese women. ... Show more

View full text

13 Citations

13 References

Related records

20 Discontinuing semaglutide after weight loss: strategy for weight maintenance and a possible new side effect

Carris, NW; Wallace, S; (...); Bunnell, B

Apr 2024 (Early Access) | CANADIAN JOURNAL OF PHYSIOLOGY AND PHARMACOLOGY

Glucagon-like peptide -1 receptor agonists (GLP-1 RAs) facilitate weight loss. Weight regain off therapy ... Show more

19 References

Free Full Text From Publisher

•••

Related records

21 Semaglutide (*Ozempic*) for Weight Loss

Apr 5 2021
|
MEDICAL LETTER ON DRUGS AND THERAPEUTICS
63 (1621`) , pp.53-54

•••

6
References

Related records

22 Effects of once-weekly semaglutide vs once-daily canagliflozin on body composition in type 2 diabetes: a substudy of the SUSTAIN 8 randomised controlled clinical trial

McCrimmon, RJ; Catarig, AM; (...); Lingvay, I

Mar 2020 | DIABETOLOGIA 63 (3) , pp.473-485

Enriched Cited References

Aims/hypothesis Intra-abdominal or visceral obesity is associated with insulin resistance and an increased ... Show more

Free Full Text From Publisher

•••

36
Citations

50
References

Related records

23 Interaction of Semaglutide and Ziprasidone in a Patient With Schizophrenia: A Case Report

Hejdak, D; Razzak, AN; (...); Jha, P
Apr 29 2024
|
CUREUS JOURNAL OF MEDICAL SCIENCE
16 (4)

Enriched Cited References

Semaglutide (Ozempic), a GLP-1 receptor agonist effective in weight management, and ziprasidone ( ... Show more

Full Text at Publisher •••

9
References

Related records

24 Subcutaneous Semaglutide Use for Weight Management: Practice

12

and Attitudes of Physicians in Israel

Dicker, D; Tamir, O; (...); Karasik, A

Oct 2023
|
ISRAEL MEDICAL ASSOCIATION JOURNAL

25 (10) , pp.644-648

Background: In 2019, 1 mg subcutaneous semaglutide was reg-istered for the treatment of diab ... Show more

9
References

Related records

25 Semaglutide-associated hyposalivation: A report of case series

Mawardi, HH; Almazrooa, SA; (...); Mawardi, MH

Dec 29 2023 | MEDICINE 102 (52)

Enriched Cited References

Rationale:Obesity and diabetes of different types are considered global health risks with rising prevalence ... Show more

Free Full Text from Publisher

17
References

Related records

26 Pharmacological profile of once-weekly injectable semaglutide for chronic weight management

Lau, DCW; Batterham, RL and le Roux, CW

Mar 4 2022
|
EXPERT REVIEW OF CLINICAL PHARMACOLOGY

15 (3) , pp.251-268

Introduction The recent approval in the USA (Food and Drug Administration), Canada (Health Canada), ... Show more

Free Full Text From Publisher

4
Citations

82
References

Related records

27 Weight loss associated with semaglutide treatment among people with HIV

Haidar, L; Crane, HM; (...); Eltonsy, S

Mar 15 2024 | AIDS 38 (4) , pp.531-535

Enriched Cited References

Objective: There is limited real-world evidence about the effectiveness of semaglutide for v ... Show more

1
Citation

15
References

Free Full Text From Publisher

•••

Related records

☐ 28

### Clinical Insight on Semaglutide for Chronic Weight Management in Adults: Patient Selection and Special Considerations

Chao, AM; Tronieri, JS; (...); Wadden, TA

2022

|

DRUG DESIGN DEVELOPMENT AND THERAPY

16 , pp.4449-4461

Losses of 5-10% or more of initial body weight are associated with improvements in obesity-related c ... Show more

Free Full Text from Publisher •••

9 Citations

93 References

Related records

☐ 29

### Weight loss response in patients with obesity treated with injectable semaglutide in a real-world setting

Alabduljabbar, K; Alsaqaaby, M; (...); le Roux, CW

Feb 2024 | ENDOCRINE 83 (2) , pp.392-398

Enriched Cited References

BackgroundObesity is defined as excess adipose tissue causing a deterioration in health, but diagn ... Show more

View full text •••

2 Citations

23 References

Related records

☐ 30

### A Real-World Study of the Effectiveness and Safety of Semaglutide for Weight Loss

Tzoulis, P; Batavanis, M and Baldeweg, S

May 2 2024

|

CUREUS JOURNAL OF MEDICAL SCIENCE

16 (5)

Enriched Cited References

Introduction Recent randomized controlled trials (RCTs) have shown the great efficacy of semaglutide in ac ... Show more

Free Full Text from Publisher •••

30 References

Related records

31 [Evaluating Weight Loss With Semaglutide in Elderly Patients With Type II Diabetes](#)

Huynh, G; Runeberg, H and Weideman, R

Feb 2023

|

JOURNAL OF PHARMACY TECHNOLOGY

39 (1) , pp.10-15

Enriched Cited References

Background: Semaglutide is an effective agent indicated for type II diabetes mellitus (T2DM) treatmen ... Show more

Free Published Article From Repository

Full Text at Publisher

•••

1 Citation

16 References

Related records

32 Effects of once-weekly semaglutide on appetite, energy intake, control of eating, food preference and body weight in subjects with obesity

Blundell, J; Finlayson, G; (...); Hjerpsted, JB

Sep 2017

| DIABETES OBESITY & METABOLISM 19 (9) , pp.1242-1251

AimThe aim of this trial was to investigate the mechanism of action for body weight loss with semaglu ... Show more

Free Full Text From Publisher

•••

253 Citations

39 References

Related records

33 Semaglutide as a promising antiobesity drug

Christou, GA; Katsiki, N; (...); Kiortsis, DN

Jun 2019 | OBESITY REVIEWS 20 (6) , pp.805-815

Semaglutide is a glucagon-like peptide-1 receptor agonist (GLP-1 RA) with a long elimination half-l ... Show more

Full Text at Publisher •••

74 Citations

95 References

Related records

34 Once-Weekly Semaglutide for Weight Management: A Clinical Review

Fornes, A; Huff, J; (...); Godfrey, M

5 Citations

43 References

Aug 2022

|

JOURNAL OF PHARMACY TECHNOLOGY

38 (4) , pp.239-246

Objective: To review the efficacy, safety, and role of the glucagon-like peptide-1 (GLP-1) receptor agonist : ... Show more

Free Published Article From Repository

Full Text at Publisher

•••

Related records

35 [An Effective Glucagon-Like Peptide-1 Receptor Agonists, Semaglutide, Improves Sarcopenic Obesity in Obese Mice by Modulating Skeletal Muscle Metabolism]

8 Citations

40 References

Ren, QJ; Chen, SC; (...); Chen, XY

2022

|

DRUG DESIGN DEVELOPMENT AND THERAPY

16 , pp.3723-3735

Purpose: This study aimed to investigate the effect of Semaglutide on skeletal muscle and its metabolomics ... Show more

Free Full Text from Publisher •••

Related records

36 Effects of Semaglutide Versus Dulaglutide on Epicardial Fat Thickness in Subjects with Type 2 Diabetes and Obesity

57 Citations

28 References

Iacobellis, G and Fricke, ACV

Apr 2020

|

JOURNAL OF THE ENDOCRINE SOCIETY

4 (4)

Background and Aims. Epicardial adipose tissue (EAT), the visceral fat depot of the heart, is a modifi: ... Show more

Free Full Text from Publisher •••

Related records

37 Efficacy and Safety of Semaglutide for Weight Loss in Obesity without Diabetes: A Systematic Review and Meta-Analysis

10 Citations

20 References

Tan, HC; Dampil, OA and Marquez, MM

Aug 2022 (Early Access)

|

JOURNAL OF THE ASEAN FEDERATION OF ENDOCRINE SOCIETIES

Background. The weight loss benefit of semaglutide in patients with diabetes is well-documentec ... Show more

Free Full Text from Publisher •••

Related records

38 The role of weight control in the management of type 2 diabetes mellitus: Perspectives on semaglutide

Kurtzhals, P; Kreiner, FF and Singh, R
Sep 2023
|
DIABETES RESEARCH AND CLINICAL PRACTICE
203

Glucagon-like peptide-1 receptor agonists (GLP-1 RAs) are widely used to address multiple aspects ... Show more

Free Full Text From Publisher

•••

1 Citation

128 References

Related records

39 Semaglutide: First Global Approval

Dhillon, S

Feb 2018 | DRUGS 78 (2) , pp.275-284

Novo Nordisk has developed a subcutaneous formulation of semaglutide (Ozempic((R))), a ... Show more

View full text •••

47 Citations

26 References

Related records

40 Sustained weight loss with semaglutide once weekly in patients without type 2 diabetes and post-bariatric treatment failure

Lautenbach, A; Kantowski, T; (...); Aberle, J

Oct 2023 | CLINICAL OBESITY 13 (5)

Enriched Cited References

About 20%-25% of patients experience weight regain (WR) or insufficient weight loss (IWL) following bε ... Show more

Free Full Text From Publisher

•••

7 Citations

26 References

Related records



41 Efficacy and Safety of Semaglutide for Weight Loss in Obesity Without

10

Diabetes: A Systematic Review and Meta-Analysis

Tan, HC; Dampil, OA and Marquez, MM

Nov 2022

|

JOURNAL OF THE ASEAN FEDERATION OF ENDOCRINE SOCIETIES

37 (2) , pp.65-72

Background. The weight loss benefit of semaglutide in patients with diabetes is well-documentec ... Show more

Free Full Text from Publisher •••

Citations

20

References

Related records

42 Semaglutide Treatment of Excessive Body Weight in Obese PCOS Patients Unresponsive to Lifestyle Programs

Carmina, E and Longo, RA

Sep 2023

| JOURNAL OF CLINICAL MEDICINE 12 (18)

Enriched Cited References

In spite of the widespread use of lifestyle modifications programs, many patients with PCOS are obese a ... Show more

Free Full Text from Publisher •••

5

Citations

27

References

Related records

43 Effect of Semaglutide on High-Fat-Diet-Induced Liver Cancer in Obese Mice

Liu, YH; Chen, SC and Zhen, RX

Jan 16 2024

| JOURNAL OF PROTEOME RESEARCH

23 (2) , pp.704-717

Enriched Cited References

This study aims to investigate the impact of semaglutide on the expression of liver cancer proteins ir ... Show more

Full Text at Publisher •••

28

References

Related records

44 Semaglutide for the treatment of obesity - a review

Bham, A and Ditta, MA

Sep 2021 | WORLD FAMILY MEDICINE 19 (9) , pp.61-64

Increasing rates of obesity and its comorbidities continue to place a burden on individuals and h ... Show more

19

References

Free Full Text From Publisher

•••

Related records

45 Efficacy and safety of semaglutide on weight loss in obese or overweight patients without diabetes: A systematic review and meta-analysis of randomized controlled trials

Gao, XQ; Hua, XL; (...); Gu, M

Sep 14 2022

| FRONTIERS IN PHARMACOLOGY 13

Objectives: This study aims to explore the weight loss effect and safety of semaglutide as a conventional ... Show more

Free Full Text from Publisher •••

20 Citations

63 References

Related records

46 Efficacy and safety of semaglutide for weight management: evidence from the STEP program

Amaro, A; Sugimoto, D and Wharton, S

Apr 14 2022 | POSTGRADUATE MEDICINE

134 , pp.5-17

Enriched Cited References

Obesity is a global health challenge. It is a multifactorial, complex, and progressive disease associate ... Show more

Free Full Text From Publisher

•••

17 Citations

77 References

Related records

47 Oral Semaglutide under Human Protocols and Doses Regulates Food Intake, Body Weight, and Glycemia in Diet-Induced Obese Mice

Rakhat, Y; Wang, L; (...); Yada, T

Sep 2023 | NUTRIENTS 15 (17)

Enriched Cited References

The first oral form of the glucagon-like peptide-1 receptor agonist, oral semaglutide, has ... Show more

Free Full Text from Publisher •••

1 Citation

24 References

Related records

© 2024 Clarivate
Training Portal
Product Support

Data Correction
Privacy Statement
Newsletter

Copyright Notice
Cookie Policy
Terms of Use

Manage cookie preferences

Follow Us

Table S2. Extracted data

| Study | | Participants | | |
|---|---|---|---|---|
| Authors and Year | Country | N | Semaglutide | Placebo |
| Gibbons et al. 2021 | UK | 13 | 13 | 13 |
| Ingersen et al. 2023 | Denmark | 31 | 16 | 15 |
| Heise et al. 2023 | Germany | 72 | 44 | 28 |
| Blundell et al. 2017 | UK | 27 | 13 | 14 |
| Hansen et al. 2024 | Denmark | 64 | 32 | 32 |
| Wilding 2021 | UK | 130 | 95 | 45 |
| Eckard (2024) | USA | 92 | 46 | 46 |
| | Total | 429 | 259 | 193 |

| Participants | | | Study design |
| --- | --- | --- | --- |
| Age (y) | | Comorbidities | 1=Parallel<br>2=Crossover |
| SEMA | PLAC | | |
| | | T2D | 2 |
| 59 (6) | 56 (6) | T2D | 1 |
| 63.7 | 60.4 | T2D | 1 |
| | | None | 2 |
| 62.7 | 63.6 | Increased fracture risk; postmenopausal women | 1 |
| 46 (13) | 47 (12) | None | 1 |
| 53 | 53 | HIV-associated lipohypertrophy | |

| Semaglutide | | Concurrent intervention | |
|---|---|---|---|
| Dose | Duration | Exercise [Y/N] | Diet [Y/N] |
| 8) and then 14 mg (weeks 8–12) | 12 weeks | N | N |
| d to 0.5 mg (weeks 4-8) until a m | l; 16 weeks | Y | N |
| weeks, 0.5 mg for 4 weeks, 1 mg | 28 weeks | N | N |
| weeks), escalating to 0.5 mg (4 w | 12 weeks | N | N |
| 5 mg to 0.5 mg after 4 weeks, an | 52 weeks | N | N |
| ery 4 weeks to reach the mainte | 68 weeks | N | N |

| Analysis method |
| --- |
| Bodpod®, Concord, CA, USA |
| DEXA |
| BOD POD measurement system |
| Bodpod®, Concord, USA |
| DXA (Hologic, Inc., Marlborough, MA, USA) |
| [illegible] subpopulation as a supportive secondary end point. |

**Table S3. Study characteristics of the included studies.**

| Study | Country | Participants (n) | Study design | Comorbidities | Duration | Semaglutide protocol | Body composition method |
|---|---|---|---|---|---|---|---|
| Blundell et al. (2017) | UK | SEMA: 13<br>PLA: 14 | Crossover | None | 12 weeks | Once weekly subcutaneous injections<br>0.25 mg for 4 weeks, then 0.5 mg for 4 weeks, then 1.0 mg for 4 weeks | ADP |
| Gibbons et al. (2021) | UK | SEMA: 13<br>PLA: 13 | Crossover | T2D | 12 weeks | Once daily oral ingestion<br>3 mg (weeks 0–4), 7 mg (weeks 4–8), 14 mg (weeks 8–12) | ADP |
| Wilding et al. (2021) | UK | SEMA: 95<br>PLA: 45 | Parallel group | None | 68 weeks | Once weekly subcutaneous injections<br>0.25 mg for 4 weeks, increasing every 4 weeks to reach 2.4 mg weekly by week 16 | DEXA |
| Ingersen et al. (2023) | Denmark | SEMA: 16<br>PLA: 15 | Parallel group | T2D | 12 weeks | Once weekly subcutaneous injections<br>0.25 mg (weeks 1–4), 0.5 mg (weeks 4–8), then 0.5 mg or 1.0 mg (weeks 9–32*) | DEXA |
| Heise et al. (2023) | Germany | SEMA: 44<br>PLA: 28 | Parallel group | T2D | 28 weeks | Once weekly subcutaneous injections<br>0.25 mg for 4 weeks, 0.5 mg for 4 weeks, then 1.0 mg for 20 weeks | ADP |
| Hansen et al. (2024) | Denmark | SEMA: 32<br>PLA: 32 | Parallel group | Increased fracture risk; postmenopausal women | 52 weeks | Once weekly subcutaneous injections<br>0.25 mg for 4 weeks, increased to 0.5 mg after 4 weeks, then to 1.0 mg after another 4 weeks | DEXA |
| Eckard et al. (2024) | USA | SEMA: 46<br>PLA: 46 | Parallel group | HIV-associated lipohypertrophy | 32 weeks | Once weekly subcutaneous injections<br>0.25 mg for 4 weeks, 0.5 mg for 4 weeks, then 1.0 mg for 24 weeks | DEXA |

SEMA = Semaglutide; PLA = Placebo; ADP = Air Displacement Plethysmography; DEXA = dual-energy x-ray absorptiometry; T2D = type 2 diabetes.
*Note: Participants supplemented with semaglutide for 20 weeks prior to initiating the aerobic exercise training program.

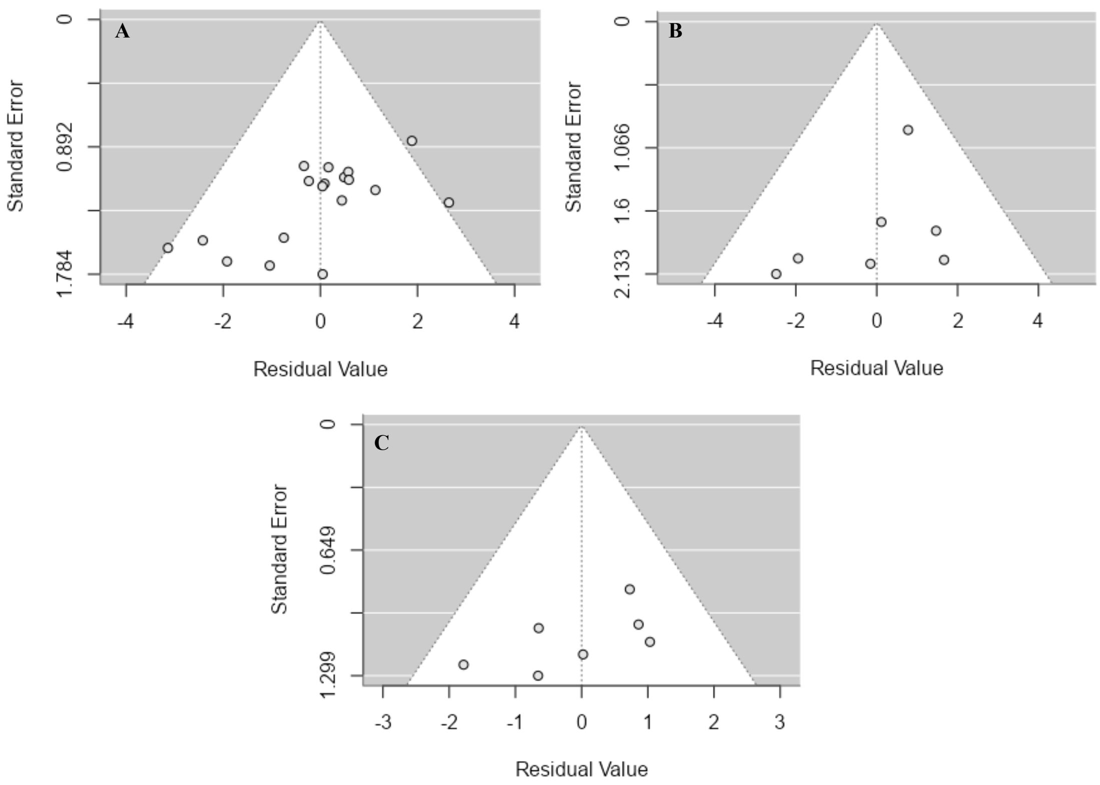


**Fig. S1.** Funnel plots of studentised residuals (x axis) by standard errors (y axis) of A) fat mass, B) fat mass, and C) fat-free mass. The white areas represent the expected confidence intervals for each calculated standard error.

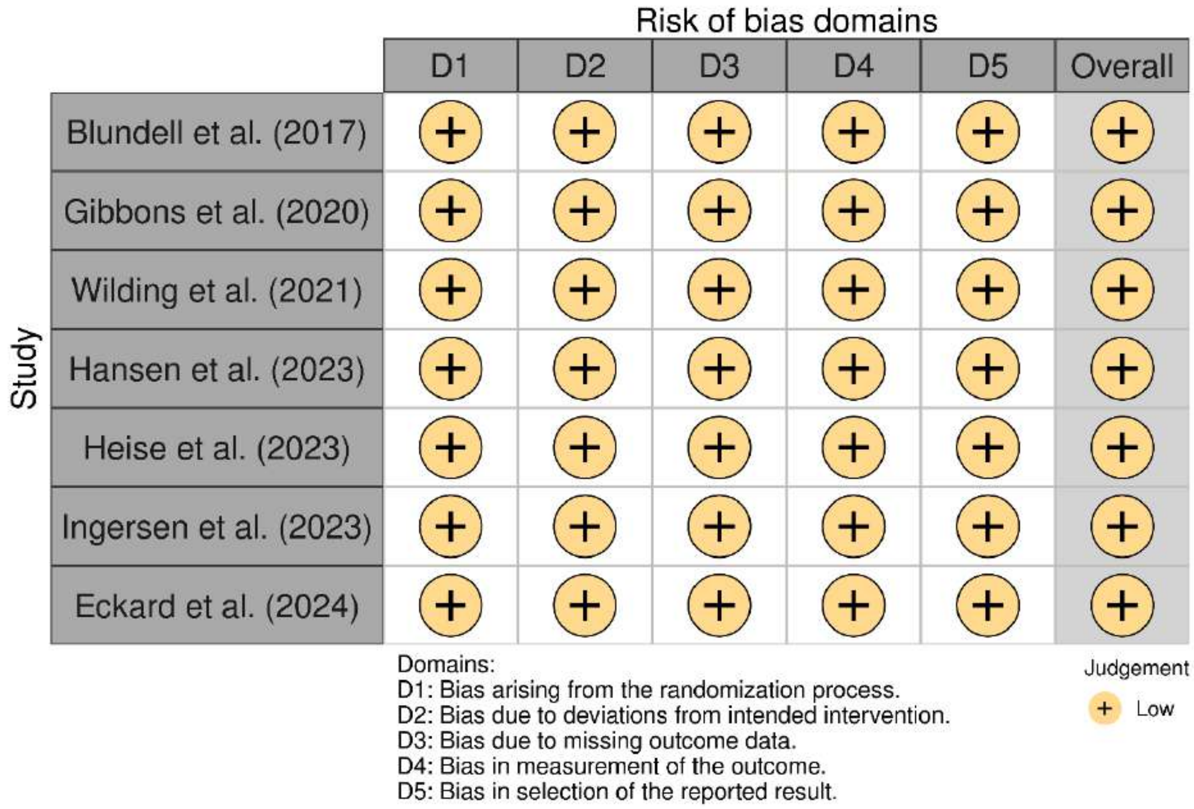


**Fig. S2.** Risk of bias assessment of the seven studies included in the meta-analysis. (Plot was created using robvis [23] and are in a colourblind-friendly colour scheme).

**Table S4.** Results of Grading of Recommendations Assessment, Development and Evaluation (GRADE)

| Analysis | GRADE items for downgrading quality of evidence | | | | | GRADE items for upgrading the quality of evidence | | | Overall* |
|---|---|---|---|---|---|---|---|---|---|
| | **Risk of bias** | **Inconsistency** | **Indirectness** | **Imprecision** | **Publication bias** | **Large effect** | **Dose-response** | **Confounding** | |
| Body mass | ↔ | ↓ | ↔ | ↔ | ↔ | ↔ | ↔ | ↔ | ⊕⊕⊕◯ Moderate |
| Fat mass | ↔ | ↓ | ↔ | ↔ | ↔ | ↔ | ↔ | ↔ | ⊕⊕⊕◯ Moderate |
| Fat-free mass | ↔ | ↓ | ↔ | ↔ | ↔ | ↔ | ↔ | -↔ | ⊕⊕⊕◯ Moderate |

* Classification based on the GRADE Handbook as: ⊕⊕⊕⊕ = High quality; ⊕⊕⊕O = Moderate quality; ⊕⊕OO = Low quality; ⊕OOO = Very low quality. ↓quality of evidence downgraded one level; ↔ no downgrade or upgrade.

**Table S5.** Deviations from registered study protocol

| **Section** | **Change** |
|---|---|
| **Title** | Changed from *"Semaglutide for weight loss: Where is the weight loss coming from? A systematic review and meta-analysis"* to *"Body Composition Changes with Semaglutide: A Systematic Review and Meta-Analysis"*. |
| **Author order** | Bryan Saunders moved from first to last author; Guilherme Giorelli became first author. |
| **Search databases** | SciELO was excluded after a preliminary search indicated few relevant results. |
| **Study design** | Initially limited to parallel-group designs, but crossover trials were also included to increase sample size. |
| **Data extraction** | An individual participant data (IPD) meta-analysis was planned, but only two studies provided IPD, making this analysis unfeasible. |
| **Data synthesis** | Meta-regressions for dosage, duration, concomitant exercise, and comorbidities were planned but not performed due to insufficient data. |
| **Timeline** | The original end date (December 2024) was substantially exceeded. |
| **GRADE assessment** | Not prespecified in the protocol but added to strengthen the conclusions. |
